# Radiomics Features of ^18^F-fluorodeoxyglucose Positron-Emission Tomography as a Novel Prognostic Signature in Colorectal Cancer

**DOI:** 10.1101/2019.12.27.19015982

**Authors:** Jeonghyun Kang, Jae-Hoon Lee, Hye Sun Lee, Eun-Suk Cho, Eun Jung Park, Seung Hyuk Baik, Kang Young Lee, Chihyun Park, Yunku Yeu, Jean R. Clemenceau, Sunho Park, Hongming Xu, Changjin Hong, Tae Hyun Hwang

## Abstract

**Purpose:** The aim of this study was to investigate the prognostic value of radiomics signatures derived from ^18^F-fluorodeoxyglucose (^18^F-FDG) positron-emission tomography (PET) in patients with colorectal cancer (CRC).

**Methods:** From April 2008 to Jan 2014, we identified CRC patients who underwent ^18^F-FDG-PET before starting any neoadjuvant treatments and surgery. Radiomics features were extracted from the primary lesions identified on ^18^F-FDG-PET. Patients were divided into a training and a validation set by random sampling. A least absolute shrinkage and selection operator (LASSO) Cox regression model was applied for prognostic signature building with progression-free survival (PFS) using the training set. Using the calculated radiomics score, a nomogram was developed, and the clinical utility of this nomogram was assessed in the validation set.

**Results:** Three-hundred-and-eight-one patients with surgically resected CRC patients (training set 228 vs. validation set 153) were included. In the training set, a radiomics signature called a rad_score was generated using two PET-derived features such as Gray Level Run Length Matrix_Long-Run Emphasis (GLRLM_LRE) and Grey-Level Zone Length Matrix_Short-Zone Low Gray-level Emphasis (GLZLM_SZLGE). Patients with a high-rad_score in the training and validation set had shorter PFS. Multivariable analysis revealed that the rad_score was an independent prognostic factor in both training and validation sets. A radiomics nomogram, developed using rad_score, nodal stage, and lymphovascular invasion, showed good performance in the calibration curve and comparable predictive power with the staging system in the validation set.

**Conclusion:** Textural features derived from ^18^F-FDG-PET images may enable more detailed stratification of prognosis in patients with CRC.

## Introduction

Colorectal cancer (CRC) is the second most common cancer in women and the third most common in men, worldwide [1]. CRC is one of the major causes of cancer-related death [2, 3]. Surgery and adjuvant chemotherapy and/or radiotherapy specific for rectal cancer are regarded as standard treatments. Currently, appropriate treatment for CRC is planned based on the results of preoperative imaging studies. When CRC is confirmed pathologically, usually via an endoscopic procedure, staging abdominopelvic computed tomography (CT) or chest CT are used next for detection of distant metastases. For rectal cancer patients, adding magnetic resonance imaging (MRI) could afford more accurate local staging, and thus could provide additional information in deciding preoperative chemoradiotherapy (preop-CRT) [4]. However, if systemic metastasis is definitely confirmed via an abdominopelvic or chest CT, adding ^18^F-fluorodeoxyglucose (^18^F-FDG) positron-emission tomography (PET) or ^18^F-FDG-PET/CT for further clinical decision is controversial. Some studies have demonstrated that conventional PET-derived parameters, such as maximum standardized uptake value (SUVmax), metabolic tumor volume (MTV), and total lesion glycolysis (TLG), carry their own significance in stratifying survival of patients with CRC [5-8]. However, other studies yielded equivocal results [9, 10]. The prognostic impact of those conventional PET-based parameters was not evident for patients with CRC, and the National Comprehensive Cancer Network guidelines recommended that ^18^F-FDG-PET or PET/CT should be used selectively for potentially surgically curable metastatic diseases or should only be used to evaluate an equivocal finding on a contrast-enhanced CT or MRI [11]. Thus, it is necessary to explore the additional clinical efficacy of PET scan in the management of CRC.

Radiomics has frequently been adopted for subtype classification, evaluation of lymph node metastasis or distant metastasis, and prediction of treatment response or prognosis in many types of cancers [12-15]. It is regarded as somewhat difficult to assess intra-tumoral heterogeneity using a single spatially-biased biopsy [16]. Detection of unseen information reflecting intra-tumoral heterogeneity might be the main motivation for applying a radiomics approach in the oncology field. Several studies have investigated the practicality of using ^18^F-FDG-PET radiomics in CRC patients. Primarily, baseline textural features, such as homogeneity, coarseness, dissimilarity and contrast from the neighborhood intensity-difference matrix, kurtosis of the absolute gradient, and coefficient of variation of SUV, have analyzed as the prognostic factors for survival and as a predictor of response to preop-CRT in patients with rectal cancer [17-20]. Nevertheless, because CRC refers to both the cancer inside the rectum (rectal cancer) and that inside the colon (colon cancer), application of baseline ^18^F-FDG-PET signatures as a prognostic factor in CRC patients who underwent curative intent resection followed by selective chemotherapy has been seldom reported.

Thus, we aimed to identify quantitative ^18^F-FDG-PET-based imaging biomarkers, using a radiomics approach, to predict survival outcomes in patients with CRC.

## Materials and methods

### Study population

We retrospectively reviewed clinical information and radiological images of patients with CRC who underwent curative-intent resection at Gangnam Severance Hospital, Yonsei University College of Medicine, between April 2008 and January 2014. Patients with CRC who underwent ^18^F-FDG-PET as a preoperative diagnostic modality were initially selected. Inclusion criteria were biopsy-proven primary CRC, ^18^F-FDG-PET used as the initial baseline diagnostic tool before starting any neoadjuvant treatment, within 2 months of surgery, and the availability of follow-up data and clinical information.

Exclusion criteria were PET images unsuitable for further analyses, PET images taken outside of our hospital, and presence of a lesion that was not identifiable or features that were not extractable from PET images. Patients whose metabolic tumor volume less than 5.0 mL were also excluded because these lesions could be affected by partial volume effects. [21, 22] A total of 381 individuals (224 men; age, 22–88 years) who had stage I-IV CRC and underwent surgery were included in this study.

This study was approved by the institutional review board of Gangnam Severance Hospital, Yonsei University College of Medicine (Seoul, Republic of Korea) (approval no. 3-2018-0177). The need to obtain informed consent was waived for this retrospective study.

### PET/CT protocol

All patients fasted for at least 6 hours before the PET/CT scan and had blood glucose levels less than 140 mg/dL before intravenous administration of ^18^F-FDG (5.5 MBq/kg of body weight). Sixty minutes after intravenous administration of ^18^F-FDG, PET/CT scans were performed with a hybrid PET/CT scanner (Biograph 40 TruePoint or Biograph mCT 64, Siemens Healthcare Solutions USA, Inc., Knoxville, TN). Whole-body CT images were obtained first for attenuation correction using automatic dose modulation with a reference of 40 mA and 120 kV without contrast enhancement. Then, PET data were acquired from the skull base to the proximal thigh for 3 minutes per bed position in three-dimensional mode. PET images from the mCT 64 scanner were reconstructed using the ordered-subset expectation maximization (OSEM) algorithm with the point spread function (PSF), time of flight (TOF) modeling, and a 5-mm Gaussian post-filtering process (21 subsets and 2 iterations). For reconstructing the PET images from the Biograph 40 scanner, only the OSEM algorithm with a 5-mm Gaussian post-filtering (8 subsets and 4 iterations) was used. Through regular standardization and quality assurance using a phantom, difference in the measurements of the standardized uptake value (SUV) between the two scanners was minimized to less than 10%.

### Feature extraction for radiomics analysis

A primary volume of interest (VOI) of ^18^F-FDG-PET was manually drawn around the tumor, avoiding physiological FDG uptake, particularly at both urinary tracts. In cases of patients with stage IV CRC, the primary lesion of the colon or rectum were considered, rather than the metastatic foci. The relevance of these manually delineated initial VOIs were assessed before performing further automatic segmentation. The final VOI of the primary tumor lesion was (semi-)automatically defined on PET images with a threshold of 40% of the SUVmax.

Forty-seven quantitative features from VOIs of each patient’s PET image were extracted using LIFEx software, which is currently an open source software (http://www.lifexsoft.org) [23]. Algorithms used to obtain histogram-based, shape and size, and second and high order features are illustrated in the Supplementary file. Conventional parameters, such as SUVmax, SUVmean, and MTV were measured from the final VOI. TLG was calculated by SUVmean x MTV.

Initially, the inter-observer agreement of the features extracted by a nuclear medicine board-certified physician and a single trained physician, both blinded to the patients’ clinical outcomes, was measured using a sample of 43 patients randomly selected from our cohort. After comparing the intraclass correlation coefficient (ICC) for these two readers, further measurements of the remaining cases were done by the single trained physician.

### Feature selection, building of rad_score, and validation

Patients were allocated to training and validation sets using random sampling at a fixed ratio; 60% of the patients were assigned to the training set and the remaining 40% were assigned to the validation set. The percentages were selected by considering the event numbers in each group.

The least absolute shrinkage and selection operator (LASSO) algorithm, in the context of a Cox model, was applied to implement a meaningful feature selection scheme, based on the association of every feature with progression-free survival (PFS) of patients in the training set [24]. Because the values obtained in the initial stage had diverse ranges, we standardized the feature values for the LASSO regression. Thus, a combination of feature variables, called the rad_score, could be generated. The steps for extracting those features used in the rad_score calculation are explained in the Supplementary file. The “glmnet” package was used to perform the LASSO Cox regression analysis.

After generating the rad_score using the LASSO COX regression model in the training set, the optimum cut-off point of the rad_score was selected on the basis of the association with PFS of patients in the training set, using X-tile software (Version 3.6.1, Yale University School of Medicine, New Haven, CT) [25]. The optimal cut-off value was defined as the value that produced the largest χ^2^ in the Mantel–Cox test and patients were divided into the high- and low-risk subgroups based on this value. This cut-off value for rad_score in the training set was applied to the validation set, to explore the potential association of the rad_score with PFS. A multivariable Cox proportional hazards model was applied to identify the potential of the rad_score as an independent prognostic biomarker in the training set, the validation set, and the overall (training + validation) set.

### Development and validation of the radiomics nomogram

To confirm the prognostic impact of using the rad_score in a clinical setting, the radiomics nomogram was generated using the training set and was then validated in the validation set. The radiomics nomogram was composed of the radiomics signature and independent clinicopathologic predictors, based on the multivariable Cox regression analysis. The Akaike information criterion (AIC) and Harrell’s concordance index (C-index) were used to compare the performance between the radiomics nomogram and the American Joint Committee on Cancer (AJCC) stage in the training set, the validation set, and the overall set.

Calibration curves, which compared the predicted survival with the actual survival, were generated to explore the performance characteristics of the developed nomogram in the validation set.

### Statistical analysis

All statistical analyses were performed using SPSS software, version 23.0 (SPSS, Chicago, IL) and R version 3.5.1 (R-project, Institute for Statistics and Mathematics, Vienna, Austria). Continuous data were described as the mean ± standard deviation and were analyzed using the Student’s t test or Wilcoxon rank sum test. Categorical data were analyzed using Pearson’s Chi square test or Fisher’s exact test for dichotomous parameters.

All radiomics features obtained from baseline PET examination were normalized by transforming the data into new scores with a mean of 0 and a standard deviation of 1. PFS was defined from the date of surgery until the date of recurrence detection, last follow-up, or death. The patients alive at last follow-up were censored. The Kaplan–Meier method was used to construct survival curves and the log-rank test was used to compare survival rates between the groups.

A univariable analysis was performed to calculate the hazard ratio of the single variables in the Cox Proportional-Hazard model after entering one of the variables under investigation in the calculation model. After calculating all hazard ratios in the univariable analysis, the parameters which showed statistical significance (*p*<0.05) or potential significance (*p*<0.1) were further used in the multivariable analysis. Multivariable survival analyses with forward stepwise selection were performed using the Cox Proportional-Hazard model to test the independent significance of different factors.

The coefficients of the multivariable Cox regression model in the training set were used to construct a nomogram with the “rms” package of R software. The calibration curves were used to evaluate the clinical usefulness of the nomogram. The AIC and C-index were calculated to compare the radiomics nomogram and AJCC stage. A smaller AIC value indicated a better goodness-of-fit for predicting outcomes and a higher C-index value indicated a better concordance of survival times [26, 27]. A two-sided *p*<0.05 was considered statistically significant.

## Results

### Patient characteristics

Three-hundred-and-eighty-one patients were included in our analysis. There were 43 recurrences after a mean follow-up period of 36.4 months (interquartile range, 27–60 months). Our initial cohort was categorized into two groups by random sampling at a fixed ratio; 228 patients (60%) were assigned to the training set and the remaining 153 patients (40%) were assigned to the validation set. There were 25 and 18 recurrences in the training and validation sets, respectively.

Patient characteristics in the training set and the validation set are shown in Table 1. Except for the lymphovascular invasion (LVI) rate, there were no statistically significant differences in clinicopathologic findings between the training set and validation set. There was no significant difference in the mean and standard deviation of rad_score between the two groups.

**Table 1.** Comparison of patients’ demographics between the training set and the validation set

|  |  | Training set | Validation set | *p* |
| --- | --- | --- | --- | --- |
|  |  | (n = 228) | (n = 153) |  |
|  |  | n (%) | n (%) |  |
| Sex | Male | 132 (57.9) | 92 (60.1) | 0.743 |
|  | Female | 96 (42.1) | 61 (39.9) |  |
| Age (years) | < 70 | 156 (68.4) | 107 (69.9) | 0.841 |
|  | ≥ 70 | 72 (31.6) | 46 (30.1) |  |
| ASA<sup>a</sup> | 1 | 110 (48.2) | 72 (47.1) | 0.163 |
|  | 2 | 88 (38.6) | 62 (40.5) |  |
|  | 3 | 30 (13.2) | 19 (12.4) |  |
| BMI<sup>b</sup> (kg/m<sup>2</sup>) | < 25 | 161 (70.6) | 115 (75.2) | 0.391 |
|  | ≥ 25 | 67 (29.4) | 38 (24.8) |  |
| Preop-CEA<sup>c</sup> (ng/mL) | < 5 | 147 (64.5) | 100 (65.4) | 0.946 |
|  | ≥ 5 | 81 (35.5) | 53 (34.6) |  |
| Tumor location | Colon | 165 (72.4) | 117 (76.5) | 0.438 |
|  | Rectum | 63 (27.6) | 36 (23.5) |  |
| Surgery type | Open | 82 (36) | 49 (32) | 0.494 |
|  | MIS<sup>d</sup> | 146 (64) | 104 (68) |  |
| Complications | No | 178 (78.1) | 127 (83) | 0.293 |
|  | Yes | 50 (21.9) | 26 (17) |  |
| Histologic grade | G1 | 23 (10.1) | 14 (9.2) | 0.936 |
|  | G2 | 186 (81.6) | 127 (83) |  |
|  | G3 and Mucinous | 19 (8.3) | 12 (7.8) |  |
| LVI<sup>e</sup> | Absent | 175 (76.8) | 98 (64.1) | 0.010 |
|  | Present | 53 (23.2) | 55 (35.9) |  |
| No. of Retrieved LNs<sup>f</sup> | (Mean ± SD) | 26.7 ± 16.7 | 25.5 ± 16.7 | 0.495 |
| LN numbers | < 12 | 25 (11) | 20 (13.1) | 0.644 |
|  | ≥ 12 | 203 (89) | 133 (86.9) |  |
| pT<sup>g</sup> | T1 – T2 | 39 (17.1) | 24 (15.7) | 0.504 |
|  | T3 | 156 (68.4) | 100 (65.4) |  |
|  | T4 | 33 (14.5) | 29 (19) |  |
| pN<sup>h</sup> | Negative | 113 (49.6) | 64 (41.8) | 0.168 |
|  | Positive | 115 (50.4) | 89 (58.2) |  |
| AJCC Stage<sup>i</sup> | I | 30 (13.2) | 24 (18) | 0.626 |
|  | II | 77 (33.8) | 46 (34.6) |  |
|  | III | 93 (40.8) | 63 (47.4) |  |
|  | IV | 28 (12.3) | 20 (13.1) |  |
| MSI<sup>j</sup> | MSS<sup>k</sup>/MSI-Low | 138 (60.5) | 102 (66.7) | 0.371 |
|  | MSI-High | 15 (6.6) | 11 (7.2) |  |
|  | No data | 75 (32.9) | 40 (26.1) |  |
| KRAS | Wild | 72 (31.6) | 53 (34.6) | 0.794 |
|  | Mutant | 35 (15.4) | 21 (13.7) |  |
|  | No data | 121 (53.1) | 79 (51.6) |  |
| Postoperative chemotherapy | No | 84 (36.8) | 61 (39.9) | 0.625 |
|  | Yes | 144 (63.2) | 92 (60.1) |  |
| Radiotherapy | No | 212 (93) | 143 (93.5) | >0.99 |
|  | Preoperative or postoperative | 16 (7) | 10 (6.5) |  |
| rad_score | (Mean ± SD) | 0.0 ± 0.2 | 0.0 ± 0.1 | 0.867<sup>l</sup> |

SD: Standard Deviation
a: American society of anesthesiology
b: Body mass index
c: Carcinoembryonic antigen
d: Minimally invasive surgery including laparoscopy and robotic surgery
e: Lymphovascular invasion
f: Lymph node
g: Patients who underwent chemoradiotherapy before surgery was ypT stage. Patients with pathologic complete response in rectal cancer was included in pT1 for statistical reason.
h: Patients who underwent chemoradiotherapy before surgery was ypN stage.

Analysis of the interobserver agreement of the features extracted from PET images yielded a mean ICC value, for the 47 radiomics features, of 0.932 (range, 0.78–0.99) (Supplementary file).

### Radiomics signature-based prediction model

Two features, Gray Level Run Length Matrix_Long-Run Emphasis (GLRLM_LRE) and Grey-Level Zone Length Matrix_Short-Zone Low Gray-level Emphasis (GLZLM_SZLGE), with coefficients of 0.07079258 and 0.11149516 respectively, were selected in the LASSO Cox regression model. The rad_score was defined as 0.07079258 × GLRLM_LRE + 0.11149516 x GLZLM_SZLGE. (Supplementary file).

The median rad_score was -0.0495 (range, -0.1394 to 1.4635). The optimum cut-off value generated by the X-tile program was 0.07 (Supplementary file). Using this value, patients were classified into a high-risk group (rad_score ≥ 0.07) and a low-risk group (rad_score < 0.07). In the training set, the rates of histological grade 3 and mucinous type were significantly higher in the high risk-group than in the low-risk group. The rate of rectal cancer in the high-risk group was marginally higher than that in the low-risk group, but the difference did not reach statistical significance (*p*=0.065). There was no difference of pT, pN, and AJCC stage between the high- and the low-risk groups (Table 2). The Kaplan–Meier curve showed that the radiomics signature was significantly associated with PFS in the training set (*p*<0.001) and in the validation set (*p*=0.008) (Fig. 1).

**Table 2.** Comparison of patients’ demographics between the high-risk group and the low-risk group in the training set

|  |  | Low-risk group | High-risk group | *p* |
| --- | --- | --- | --- | --- |
|  |  | (*n* = 195) | (*n* = 33) |  |
|  |  | *n* (%) | *n* (%) |  |
| Sex | Male | 113 (57.9) | 19 (57.6) | >0.99 |
|  | Female | 82 (42.1) | 14 (39.9) |  |
| Age (years) | < 70 | 132 (67.7) | 24 (72.7) | 0.709 |
|  | ≥ 70 | 63 (32.3) | 9 (27.3) |  |
| ASA<sup>a</sup> | 1 | 91 (46.7) | 19 (57.6) | 0.482 |
|  | 2 | 77 (39.5) | 11 (33.3) |  |
|  | 3 | 27 (13.8) | 3 (9.1) |  |
| BMI<sup>b</sup> (kg/m<sup>2</sup>) | < 25 | 132 (67.7) | 29 (87.9) | 0.032 |
|  | ≥ 25 | 63 (32.3) | 4 (12.1) |  |
| Preop-CEA<sup>c</sup> (ng/mL) | < 5 | 124 (63.6) | 23 (69.7) | 0.630 |
|  | ≥ 5 | 71 (36.4) | 10 (30.3) |  |
| Tumor location | Colon | 146 (74.9) | 19 (57.6) | 0.065 |
|  | Rectum | 49 (25.1) | 14 (42.4) |  |
| Surgery type | Open | 67 (34.4) | 15 (45.5) | 0.494 |
|  | MIS<sup>d</sup> | 128 (65.6) | 18 (54.5) |  |
| Complications | No | 155 (79.5) | 23 (69.7) | 0.303 |
|  | Yes | 40 (20.5) | 10 (30.3) |  |
| Histologic grade | G1 + G2 | 184 (94.4) | 25 (75.8) | 0.001 |
|  | G3 and Mucinous | 11 (5.6) | 8 (24.2) |  |
| LVI<sup>e</sup> | Absent | 152 (77.9) | 23 (69.7) | 0.415 |
|  | Present | 43 (22.1) | 10 (30.3) |  |
| No. of Retrieved LNs<sup>f</sup> | (Mean ± SD) | 26.8 ± 16.6 | 26.2 ± 17.3 | 0.846 |
| LN numbers | < 12 | 18 (9.2) | 7 (21.2) | 0.083 |
|  | ≥ 12 | 177 (90.8) | 26 (78.8) |  |
| pT<sup>g</sup> | T1 – T2 | 33 (16.9) | 6 (78.8) | 0.356 |
|  | T3 | 136 (69.7) | 20 (60.6) |  |
|  | T4 | 26 (13.3) | 7 (21.2) |  |
| pN<sup>h</sup> | Negative | 98 (50.3) | 15 (45.5) | 0.747 |
|  | Positive | 97 (49.7) | 18 (54.5) |  |
| AJCC Stage<sup>i</sup> | I | 26 (13.3) | 4 (12.1) | 0.386 |
|  | II | 68 (34.9) | 9 (27.3) |  |
|  | III | 80 (41) | 13 (39.4) |  |
|  | IV | 21 (10.8) | 7 (21.2) |  |
| MSI<sup>j</sup> | MSS<sup>k</sup>/MSI-Low | 117 (60) | 21 (63.6) | 0.669 |
|  | MSI-High | 14 (7.2) | 1 (3) |  |
|  | No data | 64 (32.8) | 11 (33.3) |  |
| KRAS | Wild | 63 (32.3) | 9 (27.3) | 0.829 |
|  | Mutant | 30 (15.4) | 5 (15.2) |  |
|  | No data | 102 (52.3) | 19 (57.6) |  |
| Postoperative chemotherapy | No | 73 (37.4) | 11 (33.3) | 0.797 |
|  | Yes | 122 (62.6) | 22 (66.7) |  |
| Radiotherapy | No | 187 (95.9) | 25 (75.8) | < 0.001 |
|  | Preoperative or postoperative | 8 (4.1) | 8 (24.2) |  |
| rad_score | (Mean ± SD) | 0.0 ± 0.0 | 0.3 ± 0.3 | < 0.001<sup>l</sup> |

SD: Standard Deviation
a: American society of anesthesiology
b: Body mass index
c: Carcinoembryonic antigen
d: Minimally invasive surgery including laparoscopy and robotic surgery
e: Lymphovascular invasion
f: Lymph node
g: Patients who underwent chemoradiotherapy before surgery was ypT stage. Patients with pathologic complete response in rectal cancer was included in pT1 for statistical reason.
h: Patients who underwent chemoradiotherapy before surgery was ypN stage.
i: AJCC denoted the American Joint Committee on Cancer. Patients who underwent chemoradiotherapy before surgery was yp Stage. Patients with pathologic complete response with no lymph node metastasis in rectal cancer was included in yp Stage I for statistical reason.

**Figure 1.** Kaplan–Meier plot in the training and the validation set according to the high risk and low risk group. Kaplan–Meier survival analyses according to the rad_score for patients in the training (A) and the validation set (B). The Kaplan–Meier curve showed that the radiomics signature was significantly associated with PFS in the training set (*p*<0.001) and in the validation set (*p*=0.008).

Univariable analysis demonstrated that LVI, pN, AJCC stage, and rad_score were significantly associated with PFS (all *p*<0.05) (Table 3), while complications (*p*=0.055), histological grade (*p*=0.08), and pT (*p*=0.052) showed marginal significance. Variables with *p*<0.1 in the univariable analysis (LVI, pN, AJCC stage, rad_score, complications, histological grade and pT) were altogether entered into the multivariable Cox proportional hazard model.

**Table 3.** Univariable analysis associated with the progression-free survival in the training set

|  |  | Univariable analysis |  |
| --- | --- | --- | --- |
|  |  | HR (95% CI) | *p* |
| Sex | Female | Ref |  |
|  | Male | 0.54 (0.24 – 1.2) | 0.136 |
| Age (years) | < 70 | Ref |  |
|  | ≥ 70 | 0.88 (0.38 – 2.04) | 0.773 |
| ASA<sup>a</sup> | 1 & 2 | Ref |  |
|  | 3 | 1.51 (0.44 – 5.07) | 0.505 |
| BMI<sup>b</sup> (kg/m<sup>2</sup>) | < 25 | Ref |  |
|  | ≥ 25 | 0.52 (0.19 – 1.39) | 0.193 |
| Preop-CEA<sup>c</sup> (ng/mL) | < 5 | Ref |  |
|  | ≥ 5 | 1.3 (0.58 – 2.89) | 0.52 |
| Tumor location | Colon | Ref |  |
|  | Rectum | 1.87 (0.84 – 4.12) | 0.12 |
| Surgery type | Open | Ref |  |
|  | MIS<sup>d</sup> | 1.66 (0.75 – 3.65) | 0.204 |
| Complications | No | Ref |  |
|  | Yes | 2.22 (0.98 – 5.04) | 0.055 |
| Histologic grade | G1 and G2 | Ref |  |
|  | G3 and Mucinous | 2.6 (0.89 – 7.61) | 0.08 |
| LVI<sup>e</sup> | Absent | Ref |  |
|  | Present | 3.96 (1.80 – 8.71) | < 0.001 |
| LN<sup>f</sup> numbers | < 12 | Ref |  |
|  | ≥ 12 | 0.68 (0.25 – 1.84) | 0.46 |
| pT<sup>g</sup> | T1 – T3 | Ref |  |
|  | T4 | 2.37 (0.98 – 5.67) | 0.052 |
| pN<sup>h</sup> | Negative | Ref |  |
|  | Positive | 2.95 (1.23 – 7.09) | 0.015 |
| AJCC Stage<sup>i</sup> | I & II | Ref |  |
|  | III & IV | 3.22 (1.28 – 8.1) | 0.012 |
| MSI<sup>j</sup> | MSS<sup>k</sup>/MSI-Low | Ref |  |
|  | MSI-High | 4.042e-08 (0 – Inf) | 0.997 |
|  | No data | 1.25 (0.57 – 2.77) | 0.571 |
| KRAS | Wild | Ref |  |
|  | Mutant | 1.84 (0.41 – 8.25) | 0.424 |
|  | No data | 1.57 (0.52 – 4.75) | 0.419 |
| Postoperative chemotherapy | No | Ref |  |
|  | Yes | 0.84 (0.36 – 1.97) | 0.7 |
| rad_score<sup>l</sup> | Continuous | 4.91 (1.73 – 13.92) | 0.002 |

HR: Hazard Ratio, CI: Confidence Interval
SD: Standard Deviation
a: American society of anesthesiology
b: Body mass index
c: Carcinoembryonic antigen
d: Minimally invasive surgery including laparoscopy and robotic surgery
e: Lymphovascular invasion
f: Lymph node
g: Patients who underwent chemoradiotherapy before surgery was ypT stage. Patients with pathologic complete response in rectal cancer was included in pT1 for statistical reason.

In the multivariable analysis with forward selection, presence of LVI (hazard ratio [HR] 3.73; 95% CI, 1.63–8.47; *p*=0.001), pathological node-positivity (HR 2.52; 95% CI, 1.01–6.26; *p*=0.046), and a high rad_score (HR 7.82; 95% CI, 2.36–25.85; *p*<0.001) remained independent predictors of a worse prognosis in the training set. The rad_score was also identified as an independent prognostic factor in the validation and the overall set (Table 4).

**Table 4.** Multivariable analysis associated with progression-free survival in the training set, validation set and overall set

|  |  | Training set |  | Validation set |  | Overall set |  |
| --- | --- | --- | --- | --- | --- | --- | --- |
|  |  | HR (95% CI) | *p* | HR (95% CI) | *p* | HR (95% CI) | *p* |
| LVI<sup>a</sup> | Absent | Ref |  |  |  | Ref |  |
|  | Present | 3.73 (1.64 – 8.47) | 0.001 |  |  | 2.37 (1.22 – 4.59) | 0.010 |
| pT<sup>b</sup> | T1 - T3 |  |  | Ref |  | Ref |  |
|  | T4 |  |  | 4.33 (1.66 – 11.29) | 0.002 | 2.22 (1.16 – 4.25) | 0.016 |
| pN<sup>c</sup> | negative | Ref |  | Ref |  | Ref |  |
|  | positive | 2.52 (1.01 – 6.26) | 0.046 | 3.38 (0.96 – 11.85) | 0.056 | 2.24 (1.05 – 4.80) | 0.037 |
| rad_score<sub>d</sub> |  | 7.82 (2.36 – 25.85) | <0.001 | 12.18 (2.21 – 66.90) | 0.004 | 8.47 (3.21 – 22.34) | <0.001 |

HR: Hazard Ratio, CI: Confidence Interval
a: lymphovascular invasion
b: Patients who underwent chemoradiotherapy before surgery was ypT stage. Patients with pathologic complete response in rectal cancer was included in pT1 for statistical reason.
c: Patients who underwent chemoradiotherapy before surgery was ypN stage.
d: continuous variable

### Calibration and discriminative performance measurement of radiomics nomogram

A radiomics nomogram was established using three variables selected in the stepwise multivariable analysis in the training set (Fig. 2). Table 5 shows the comparison of the C-index and AIC between the radiomics nomogram and the AJCC stage. In the validation set, the radiomics nomogram showed higher fitness for AIC than that in the AJCC stage (154.19 in model 4 vs. 156.861 in model 3). Radiomics nomogram and the AJCC stage showed similar predictive power of C-index after 1000 times bootstrapping of the data (0.715; 95% CI [0.561-0.874] in model 4 vs. 0.62; 95% CI [0.516-0.705] in model 3; *p*=0.101).

**Table 5.**
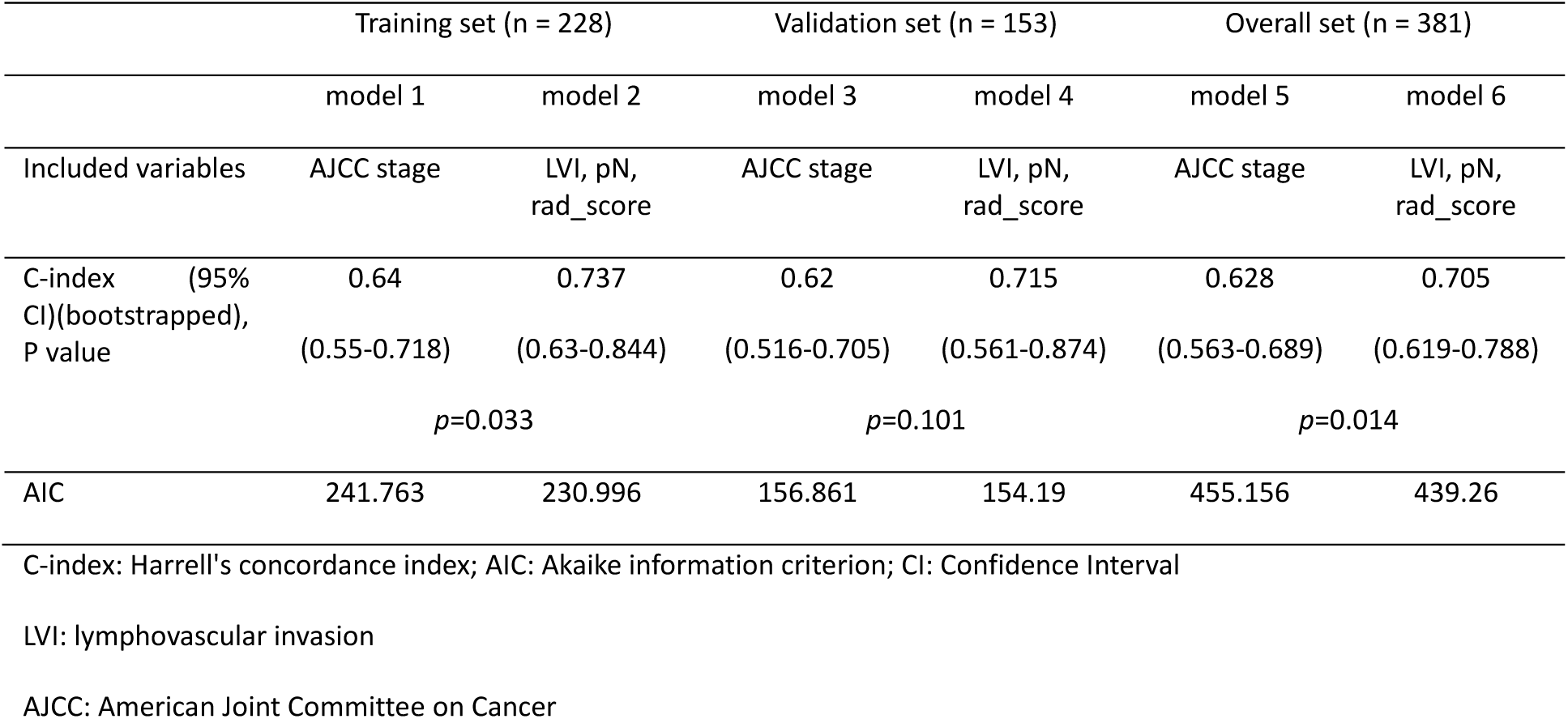
Comparison of C-index and AIC between radiomics nomogram and AJCC stage in the training, validation, and overall set

**Figure 2.** Developed nomogram to predict survival using the training set. Drawing a vertical line to the points’ axis from specific variable could determine how many points toward the probability of PFS the patient receives. The process was repeated for each variable such as LVI, N stage and rad_score, and the points for each of the risk factors were added. The final total was then located on the Total Points axis.

The calibration curve of the radiomics nomogram for estimating PFS showed good agreement between prediction and observation in the validation set (Fig. 3).

**Figure 3.** Calibration curve of radiomics nomogram in the validation set. Calibration curves for the radiomics nomograms of PFS in the validation set for agreement between the estimated and the observed 1-, 2-, and 3-year PFS. Nomogram-estimated PFS is plotted on the x-axis; the observed PFS is plotted on the y-axis. The validation set was divided into two groups according to the survival duration; nomogram-estimated PFS and observed PFS based on the 1-, 2-, and 3-year PFS were calculated in each group. The diagonal dotted line represents the optimal estimation of PFS by an ideal model, in which the estimated outcome perfectly corresponds to the actual outcome. The solid lines with black, gray, and blue colors represent the performance of the nomogram. A close alignment with the diagonal dotted line indicates better estimation. PFS, progression-free survival

### Comparison of survival within the same stages based on the rad_score

Kaplan-Meier curves for the subgroups stratified according to the radiomics signature (high-risk group versus low-risk group) revealed significantly poor PFS of high-risk groups in stage II (*p*=0.031) and stage III patients (*p*<0.0001) respectively (Fig. 4).

**Figure 4.** Comparison of progression-free survival of the high- and low-risk groups defined by radiomics signature according to the stages in the overall (training + validation) set. Kaplan–Meier survival analyses according to the radiomics signature (high-risk group versus low-risk group) in the stage II (A), stage III (B) and stage IV patients (C). The Kaplan–Meier curve showed that the radiomics signature was significantly associated with PFS in the stage II (*p*=0.031) and in the stage III patients (*p*<0.0001) respectively.

## Discussion

In the present study, we developed and validated a radiomics nomogram using preoperative ^18^F-FDG-PET images for personalized prediction of PFS in patients with CRC. This study demonstrated that the features derived from baseline PET could help stratify patients’ prognosis and the radiomics nomogram showed comparable performance to AJCC stage for predicting PFS in patients with CRC.

Radiomics analysis with PET has been recently investigated for patients with CRC. Bundschuh et al. reported that the coefficient of variation of SUV correlated with PFS [17]. Bang and colleagues demonstrated that textural features, including kurtosis of the absolute gradient, measured before starting preop-CRT, was associated with 3-year disease-free survival (DFS) [18]. Lovinfosse and colleagues revealed that homogeneity and coarseness were associated with DFS and dissimilarity and contrast from the neighborhood intensity-difference matrix were correlated with overall survival, respectively [19]. Nonetheless, previous studies investigating the impact of radiomics derived from PET mainly focused on patients with rectal cancer, who underwent preop-CRT, and included a relatively small number of patients and no validation set [17-19]. The impact of PET-based signature analysis in patients undergoing curative-intent radical surgery, with or without chemotherapy, has not been extensively investigated to date.

A radiomics signature-based nomogram has been developed and validated for preoperative prediction of survival in breast cancer, gastric cancer, and early-stage non-small cell lung cancer patients [28-30]. In our study, the developed nomogram incorporated a radiomics score derived from two components of the PET-based features, pN and LVI. Both are well-known clinicopathological prognostic factors in CRC patients [31-33]. Unexpectedly, AJCC stage was not identified as a significant factor in our multivariable COX proportional hazard model. Our study included patients who underwent surgeries first, with curative-intent, even in patients diagnosed as stage IV. Therefore, overall prognosis of stage IV patients in this study might be better than that of commonly reported stage IV groups that included initially unresectable patients, which might be the main reason for the greater prognostic power of node-positivity than stage itself, considering multicollinearity.

The radiomics signature, which was used to divide the patients into high-risk and low-risk groups, was not related to pT, pN, and AJCC stage in the training set. In addition, the patient’s PFS in stage II and stage III CRC could be stratified according to the radiomics signature. Collectively, these findings suggested that the radiomics signature might have different uses, compared to clinicopathological parameters, in predicting prognosis of patients with CRC, which may be an advantage in applying this image biomarker. In the current clinical setting, estimating prognosis and using chemotherapy with appropriate selection of chemotherapy agents are mainly dependent on the postoperative pathological stages. Wide variations in survival among patients with the same AJCC stage are already well-known in patients with CRC [34, 35]. Prognostic biomarkers may help to select patients at high risk of recurrence, allowing customization of the follow-up monitoring process for the individual patient. The detection of circulating tumor (ct) cells or ctDNA after surgery may provide good sensitivity and specificity, although such a test would have to be compared with CEA-based detection of tumor recurrence [36-39]. Patients with a high risk of recurrence may be good candidates for liquid biopsy-based follow-up; this would be a more reasonable approach in the context of medical resource use, considering the additional cost imposed by liquid biopsies. Preoperatively, neoadjuvant chemotherapy prior to surgery has been investigated as an option for treatment, especially for locally advanced, operable colon cancer patients [40, 41]. In this context, proper risk stratification prior to definite surgery may be essential for selecting candidates for neoadjuvant chemotherapy. Our study demonstrated that PET-based radiomics analysis could enhance patient stratification and thus could be a potential way for guiding personalized care before or after surgery in CRC patients, although the contribution of this approach would need to be validated in future.

There were several limitations in the present study. Placement of the VOI was done by manual selection of the entire lesion, which was a time-consuming and labor-intensive task in some cases. Although our study revealed that combination of two baseline features derived from PET could be used as a useful image biomarker, the biological underpinnings of this correlation were not evident [42]. Moreover, previously identified PET-derived radiomics features, such as coefficient of variation of SUV, kurtosis of the absolute gradient, homogeneity and coarseness, and the dissimilarity and contrast from the neighborhood intensity-difference matrix, which were proven to be important prognostic factors especially in patients with rectal cancer, were not reproducible in our radiomics signatures. [17-19] The discordance of the results across studies might not be easily understood and the explanation is likely to be multifactorial. First, various computer algorithms have been used for feature extraction, and the types of features extracted by each algorithm were not uniform. Moreover, we applied standardization of features as a preprocessing step before entering the data into the LASSO COX model, following a method that has already been adopted for radiomics analysis in patients with breast or gastric cancer [28, 30]. However, most previous studies of PET-derived radiomics analysis in patients with CRC did not use standardization as a preprocessing step [17-19, 43]. The effect of this preprocessing needs to be investigated with more clinical data. Second, accurate tumor segmentation remains challenging, and there is no standard or recommended method even for radiomics analysis of PET, although the ICC value was relatively high in our study. Third, several studies attempted to determine relatively robust features that could minimize the radiomics feature variations using PET imaging [44-46]. Although robust feature identification and standardization might be essential, the results of those studies are also somewhat inconsistent. Different settings for evaluation of the radiomics features, such as using phantom versus real patients’ data, absence of features with the same names, and different sources of variability and statistical assessment of robustness, make it difficult to compare the results between the studies [46]. Finally, the differences in imaging protocols of different PET/CT systems might potentially hinder the robustness of feature extractions. Therefore, for greater acceptance of radiomics in clinical decision making, correlation and correspondence among different imaging protocol is an important step. We considered that all of these situations might be common limitations in the overall application of PET-derived radiomics in the oncology field. Due to the retrospective design of the study, genomic data, such as MSI or KRAS data, were not available for all patients and thus could not be included in the multivariable analysis as an important genetic biomarker. However, the prognostic or predictive value of these genetic mutations in CRC have previously shown some contradictory results [47]. It may be difficult to generalize our results because the study was performed in a single institution, included a relatively small number of patients, and used only an internal validation set. In addition, although our radiomics nomogram showed good calibration in the validation set, this did not improve the predictive accuracy for survival as compared with the AJCC stage. Thus, we believe that our newly developed radiomics nomogram cannot replace the currently used staging-based strategy; however, our radiomics nomogram can be used to effectively discriminate patients’ survival within the same stage of CRC. Although further studies may be required to confirm our results, this study could be used as a proof-of concept of the potential use of this approach in clinical practice.

In conclusion, texture analysis from PET images yield prognostic information about PFS in CRC patients. Assessing these radiomics data during the baseline diagnostic stage may assist in predicting long-term outcomes. The reproducibility of our feature-based prediction model should be evaluated in further large-scale studies.

## Data Availability

The datasets generated and/or analyzed during the current study are available from the corresponding author on reasonable request pending the approval of the institution(s) and trial/study investigators who contributed to the dataset.

## Acknowledgement

We would like to thank Editage (www.editage.co.kr) for English language editing.

